# The social determinants of suicide: an umbrella review

**DOI:** 10.1101/2024.08.09.24311718

**Authors:** Gallagher Kerrie, Phillips Grace, Corcoran Paul, Platt Stephen, McClelland Heather, O’ Driscoll Michelle, Griffin Eve

## Abstract

Previous research has highlighted the role of social determinants of health on mental health outcomes, but their impact on suicide mortality is less understood. The aim of this umbrella review was to systematically examine the association between 10 social determinants of health, as defined by the World Health Organization, and suicide mortality. A keyword search of titles and abstracts was conducted in six digital databases for studies published to 24 August 2023. Inclusion criteria were peer-reviewed systematic reviews and meta-analyses in English examining the association between these determinants and suicide. Methodological quality was assessed using an adapted AMSTAR-2 tool. Due to significant heterogeneity in the included studies, a meta-analysis was not undertaken. A narrative synthesis, structured by social determinant, was conducted. 49 records (25 meta-analyses and 24 systematic reviews) were eligible for inclusion in this review. The social determinants with the most available evidence were housing, basic amenities and the environment (*n*=21), income and social protection (*n*=13), unemployment (*n*=8) and early childhood development (*n*=6). Limited evidence was identified for education (*n*=3), social inclusion and non-discrimination (*n*=3) and working life conditions (*n*=3). No reviews examined the relationship between affordable healthcare services, structural conflict or food insecurity and suicide mortality. There was evidence of a modest effect of social determinants on suicide mortality. Most evidence related to unemployment, job insecurity, income and social protection and childhood adversity. The methodological quality of the included reviews varied considerably. High-quality research fully exploring the relationship between social and environmental factors and suicide risk is needed.

## INTRODUCTION

The complex and multi-faceted nature of suicide is widely recognised in multi-level, interdisciplinary models of the behaviour, that typically conceptualise suicide risk as an interaction between distal (predisposing) factors (e.g., early life adversity, genetics, family history) and proximal (precipitating) factors (e.g., recent adverse life events, psychopathology) (1,2). Turecki and colleagues propose a biopsychosocial model which adds a third (developmental) set of factors, which include chronic substance misuse and personality traits. “*Sociological, demographic, economic and environmental factors may influence any or all of the distal, developmental and proximal factors*” (3). In particular, socioeconomic factors appear to have a strong association with suicide, particularly among men (4).

In recent times, the broader environmental and social context in which suicidal behaviour occurs has received more attention from suicide researchers and in national suicide prevention strategies and action plans. These social determinants of health, defined as “*the conditions in which people are born, grow, work, live, and age, and the wider set of forces and systems shaping the conditions of daily life*”, including socioeconomic context, inequalities, poverty, environmental factors and healthcare access (1), can be addressed across multiple population levels, ranging from global interventions (via collaborative, multi-sectoral action at national, regional and local levels), to targeted, person-centred approaches. A greater focus on the social determinants of suicide may help to identify opportunities to inform broader inter-governmental policies (5). Such guidance would help to target suicide prevention strategies and encourage more upstream measures to tackle health inequalities (6).

Previous research has identified the role of social determinants in the aetiology of mental ill-health. For example, an umbrella review of interventions addressing the social determinants of mental health found that welfare benefits in particular may reduce socioeconomic inequalities associated with mental health outcomes (7). However, to date there has been no systematic review which has examined the social determinants of health associated specifically with suicide mortality. The aim of this umbrella review was to fill this important knowledge gap.

## METHODS

### Search strategy and eligibility criteria

This umbrella review as based on research evidence obtained from systematic reviews and meta-analyses. The review was registered with PROSPERO (ID: CRD42023447175; 31 July 2023) and follows the PRISMA Guidelines for Systematic Reviews (8). A keyword search of titles and abstracts in six digital databases (PubMed, CINAHL, Web of Science, Scopus, PsycINFO and Embase) was conducted for studies published to 24 August 2023 (see Supplementary File 1). Individual keyword searches were constructed for 10 social determinants, as defined by the World Health Organisation (1). These included: income and social protection; education; unemployment and job security; working life conditions; food insecurity; housing, basic amenities and the environment; early childhood development; social inclusion and non-discrimination; structural conflict; and access to affordable health services of decent quality.

Inclusion criteria were peer-reviewed systematic reviews and meta-analyses which examined the association between any social determinant of health (as defined above) and suicide mortality, written in the English language. Primary research studies, conferences abstracts, reports, book chapters and dissertations were excluded. If a study examined multiple suicide-related outcomes, we only included the data specifically relating to suicide mortality. Studies were excluded when suicide mortality data could not be separated from other suicide-related outcomes (e.g., ideation, suicidal behaviour, suicide attempt). Similarly, any review which covered only one study relating to suicide mortality was excluded. A list of studies which were excluded on this basis can be found in Supplementary File 2.

The titles and abstracts were independently screened by two researchers (KG & GP) in Covidence. Disagreement was resolved through discussion among the pair or by consulting a third researcher (PC, SP, HMC OR EG). Articles included for full-text screening were assessed against the inclusion criteria by two (KG & GP) researchers. The reference lists of included studies were hand searched by two authors (KG & GP) for further potentially eligible studies; any eligible studies were uploaded to Covidence for duplicate independent screening.

### Data extraction

Data extraction was conducted using a pre-developed data extraction form (Supplementary File 3), which was initially piloted on three of the included studies. For each eligible study, we extracted data on the number and characteristics of primary studies, effect sizes and 95% confidence intervals, qualitative findings, and quality appraisal. Data were independently extracted by KG, GP and HMC and were cross-checked by one reviewer (KG & GP) to ensure reliability. Any discrepancies were resolved through discussion.

The methodological quality of included studies was critically appraised using an adapted version of the AMSTAR-2 tool (9), which was specially developed for application in the assessment of systematic reviews and meta-analysis. The tool covers 16 criteria, including study selection and screening, data extraction and synthesis, as well risk of bias assessment. Each criterion is rated as “*yes*,” “*partially yes*,” or “*no*.” The quality of included reviews/meta-analyses is rated as high (zero or one non-critical weakness), moderate (more than one non-critical weakness), low (one critical flaw with or without non-critical weaknesses) and critically low (more than one critical flaw with or without non-critical weaknesses). As the AMSTAR 2 is only partially applicable for non-intervention systematic reviews, we adapted one critical question: ‘*Did the review authors provide a list of excluded studies and justify the exclusions?*’ (10). If the authors included reasons for exclusions in their PRISMA flow diagram, we answered, ‘*partial yes’*, rather than ‘*no*’. We excluded one non-critical question, ‘*Did the review authors report on the sources of funding for the studies included in the review?*’. All studies were assessed by KG, GP and HMC independently and cross-checked; any disagreements were resolved though discussion. We provided a global rating of methodological quality (critically low, low, moderate, or high), as recommended. An adapted version of the tool can be found in Supplementary File 4.

Results of all eligible systematic reviews and meta-analyses, regardless of topic overlap, were included in the review (11). Study overlap was identified and managed by two authors (KG & GP) and documented in an excel file. Each overlapping study was colour coded and tagged to identify overlap between primary studies in the included reviews. Using guidance from the Cochrane Handbook for Overviews of Reviews, we decided not exclude studies solely based on duplication of primary studies across different reviews (11). There were several reasons for taking this approach. First, as research on the social determinants of suicide is relatively new and evolving, we aimed to provide a comprehensive overview of the existing evidence. Moreover, it is plausible that the systematic reviews and meta-analyses identified in our overview offer unique perspectives and differing methodological approaches. By including all relevant reviews regardless of overlap in primary studies, we aimed to capture diverse interpretations and synthesis of the evidence, thereby enriching the breadth and depth of our overview. Furthermore, we decided not to exclude reviews based on poor methodological quality. This decision was taken as some reviews may have identified key primary studies that are central to understanding the nuances of the topic area. By adopting a more inclusive approach, we hoped to mitigate the risk of overlooking important evidence and ensure a more balanced representation of the literature.

### Data synthesis

Due to the heterogeneity of included studies in respect of methods, outcome measures, topic areas and contexts, a meta-analysis was not undertaken. A narrative synthesis is provided, grouping reviews into topic areas. Meta-analysis results from the individual reviews were summarised visually using Stata Version 17. Odds ratios, relative risks, risk ratios, incidence rate ratios, hazard ratios and standardised mortality ratios were considered equal measures of risk due to the low rate of suicide mortality in the general population (rare disease assumption).

## RESULTS

The initial database search yielded 9,913 records. An additional record was identified from hand searching reference lists. Following the elimination of duplicates (*n*=5,206), 4,707 unique records underwent screening against the inclusion criteria at the title and abstract stage. Of these, 229 (plus the additional record) proceeded to full-text screening and 181 were excluded (see Figure 1 for reasons). Ultimately, 49 records, 25 meta-analyses and 24 systematic reviews met our inclusion criteria. Figure 1 outlines the flow of information in the different phases of this umbrella review.

**Figure 1:**
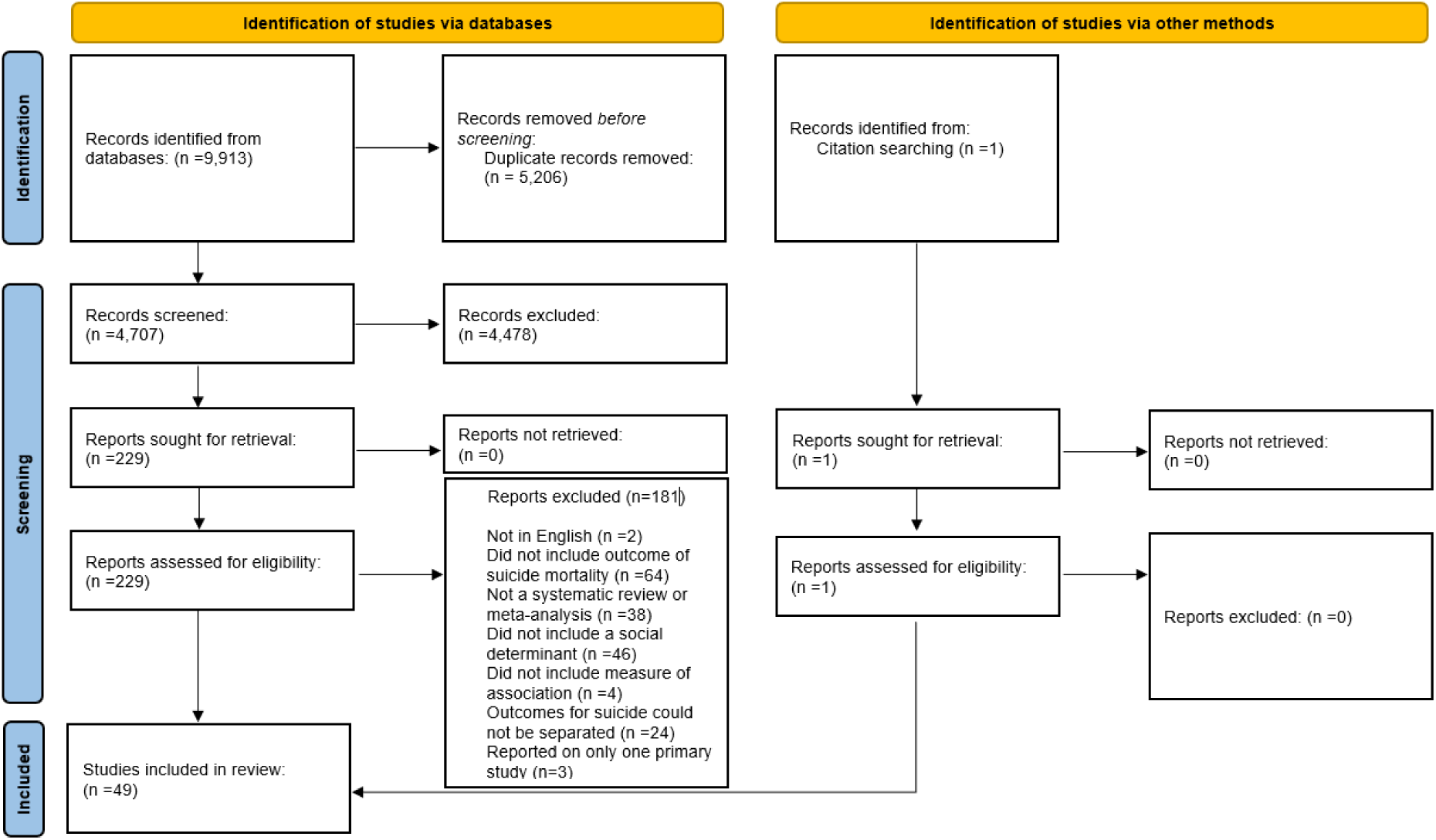
PRISMA 2020 flow diagram for new systematic reviews which included searches of databases and other sources.

All social determinants explored by each review in relation to suicide mortality were included in this umbrella review. Data extracted from eligible studies included: the determinant(s) of interest, participant demographics, number of studies related to suicide mortality and results. The years of primary studies in each review ranged from 1970 to 2021, and all reviews were published between 2004 and 2023. The included articles addressed seven of the ten social determinants examined. The social determinants with the most available evidence were housing, basic amenities and the environment (*n*=21), income and social protection (*n*=13), unemployment (*n*=8) and early childhood development (*n*=6). Limited evidence was identified for education (*n*=3), social inclusion and non-discrimination (*n*=3) and working life conditions (*n*=3). Notably, we did not identify any reviews that examined the relationship between affordable healthcare service of decent quality, structural conflict or food insecurity and suicide mortality. The characteristics of the systematic reviews and meta-analyses included in this review are detailed in each of the results tables.

### Quality of the included reviews

In this umbrella review, 25 studies were appraised as critically low and 13 were categorised as low quality. Within the critical domains of the AMSTAR-2 tool, many studies rated critically low/low for the following reasons: (i) failure to register a protocol prior to conducting the review (*n*=21); (ii) inadequate assessment of bias in the primary studies included (*n*= 19); (iii) insufficient discussion of the impact bias when interpreting the results of the review (*n*= 26) and; (iv) failure to assess publication bias, where applicable (*n*=6) (9). Four studies received a moderate assessment and the remaining seven were rated as high quality. The global ratings of systematic reviews and meta-analyses included is this review are outlined in Table 1.

**Table 1:**
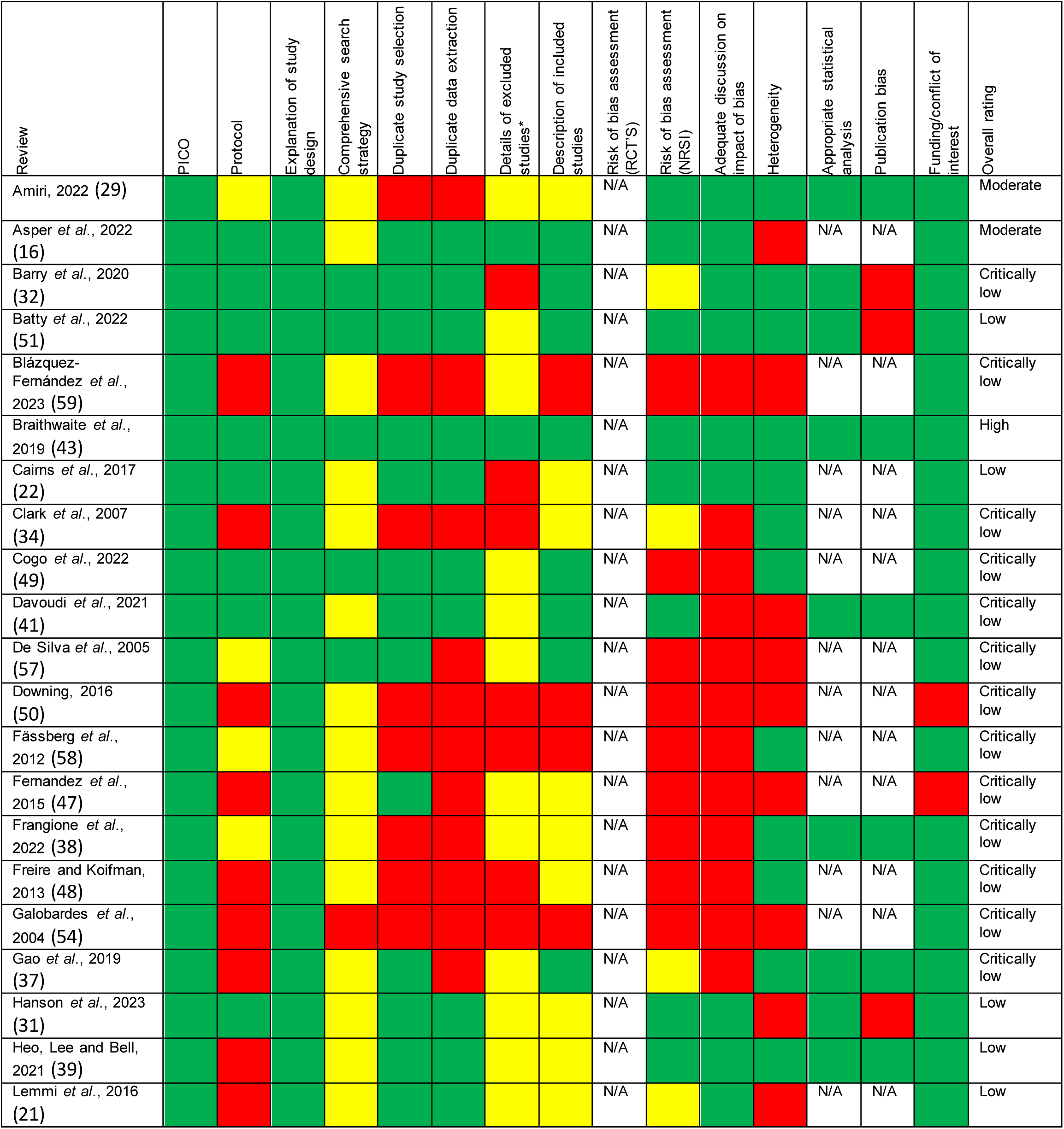
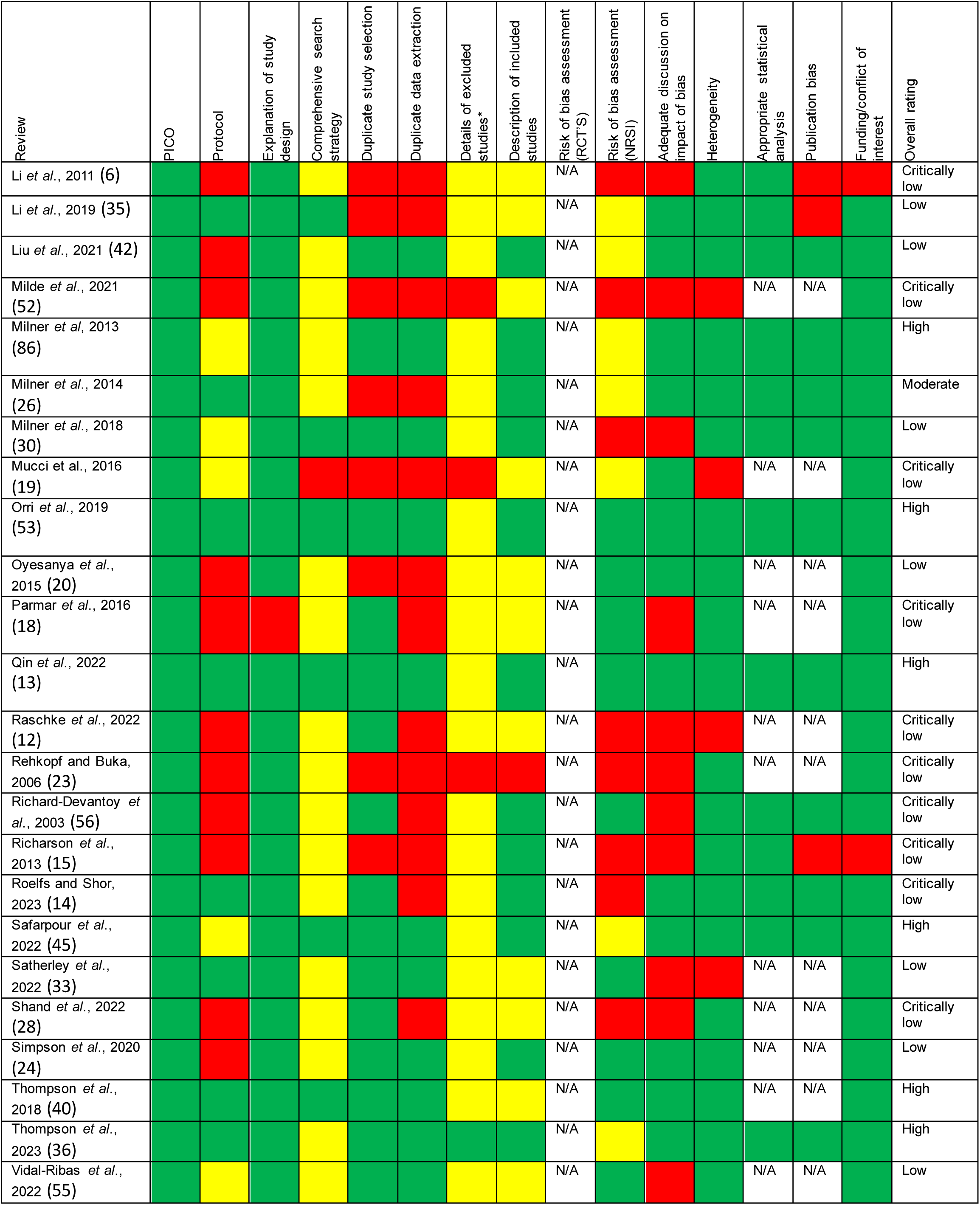
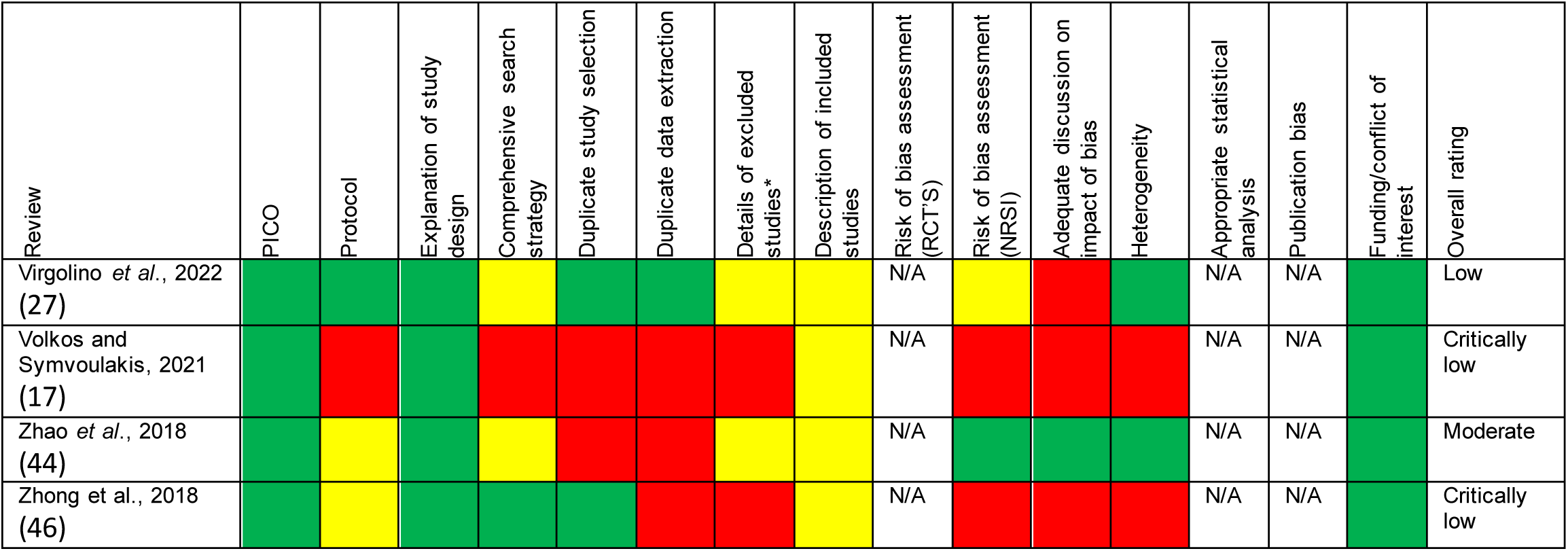
Global ratings of systematic reviews and meta-analyses from adapted AMSTAR-2 tool.

### Income and social protection (n=13)

The four meta-analyses (see figure 2) and nine systematic reviews included in this domain covered several topics, including: low income (6,12,13), financial stress (14), debt (15), economic recession (16–20), poverty (21), socio-economic disadvantage (22,23) and social security policy (24) (see Table 2). Three systematic reviews found an association between low income and the risk of suicide (6,12,13).

**Figure 2:**
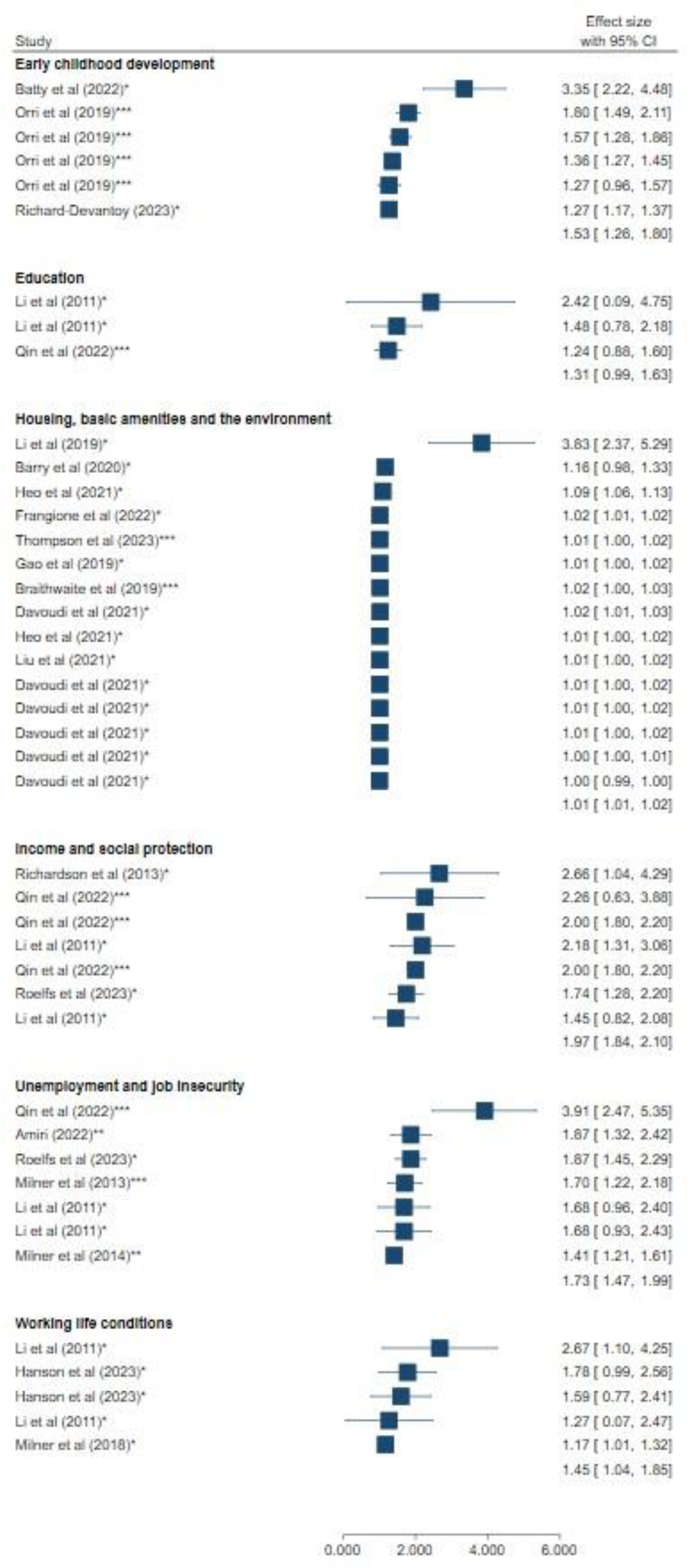
Forest plot visualisation of meta-analyses results for social determinants of suicide.

**Table 2:** Characteristics of studies relating to income and social protection.

| Review | Aim | Population(s) of interest | No. of studies related to suicide mortality | Types of included studies | Year range of included studies | Countries included | Main Findings |
| --- | --- | --- | --- | --- | --- | --- | --- |
| Roelfs *et al.*, 2023 (14)<br>Meta-analysis | To determine the suicide risk following unemployment or financial stress | Individuals with mental illness | n=23 | Case-control, cohort | 2000-2019 | Denmark, England, India, Hong Kong, New Zealand, US, Taiwan, Italy, Estonia, Germany, Turkey, Canada, Sweden | The financially stressed were 74.0% more likely to die by suicide (95% CI 36.8%, 121.5%).<br>Subgroup analyses showed that the relative suicide risk was lower than for studies that controlled for baseline physical health (47.0% elevated risk) and slightly higher than for studies that controlled for baseline mental health (78.4% elevated risk).<br>Neither were statistically significant.<br>The authors used sub-group analysis to assess the impact of bias. |
| Qin *et al.*, 2022 (13)<br>Meta-analysis | To provide an overview of research on this topic, to determine the prevalence of socioeconomic, psychiatric and physical health risk factors in suicide decedents of middle age, and to present the pooled risk of suicide conferred by these factors at midlife | Suicide decedents | n=3 | Cross-sectional, cohort | 2003-2018 | New Zealand, Hong Kong, Sweden | Low income was associated with a 2.26 (95% CI 1.16, 4.41) relative risk of suicide.<br>In a sub-group analysis by gender the relative risk for males was 2.00 (95% CI 1.81, 2.21) and 1.71 (95% CI 1.46, 1.99) for females.<br>All three studies were high quality. |
| Richardson *et al.*, 2013 (15)<br>Meta-analysis | To systematically review the relationship between personal unsecured debt and health | Individuals with unsecured debt | n=4 | Psychological autopsy studies (case-controlled) | 2006-2010 | China | The results showed a strong relationship between suicide and debt, with suicide being associated with having nearly an eight-fold risk of debt.<br>Risk of bias was not assessed. |
| Li *et al.*, 2011 (6)<br>Meta-analysis | To systematically review (a) the risk of suicide associated with high prevalence disorders and key SES measures in population-based studies of suicide; and (b) estimates the population attributable risk of suicide associated with psychiatric disorders and SES measures | Not reported | n=4 | Case-control, cohort | 1994-2003 | Denmark, Canada, Sweden, Finland, New Zealand, Lithuania | The pooled relative risk for suicide associated with low income was 2.18 (95% CI 1.47, 3.22) and 1.45 (95% CI 0.95, 2.21) for males and females, respectively.<br>The quality of the included studies was not reported. |
| Asper *et al.*, 2022 (18)<br>Systematic review | To assess the impact of the COVID-19 pandemic, previous pandemics | General population | n=37 | Longitudinal cohort, repeated cross-sectional | 2010-2020 | Italy, Greece, Spain, the European Union, Canada, England, the USA and South Korea | 19 studies found increased suicide rates at the level of the total population after the start of the 2008 economic crisis.<br>Some studies reported increases in suicide rates in specific population subgroups, among men or attributable to specific factors such as unemployment.<br>Studies relating to suicide and the economic crisis were moderate to high quality. |
| Raschke *et al.*, 2022 (12)<br>Systematic review | To investigate socioeconomic risk factors for suicidal behaviours (suicidal ideation, attempted suicides, and completed suicides) in South Korea | General population | n=6 | Prospective cohort, retrospective cohort, case-control | 2006-2019 | South Korea | Low income is significantly associated with an increased risk of suicide.<br>Outcomes were controlled for covariates such as marital status, area of residence, and age.<br>Risk of bias was not assessed. |
| Simpson *et al.*, 2021 (24)<br>Systematic review | To provide a synthesis of observational literature on the effects on mental health and inequalities in mental health of social security reforms | Adults aged 25 years and over | n=4 | Longitudinal studies | 2016-2020 | United Kingdom, United States | Suicide rates generally declined when measures were introduced to safeguard income (retirement benefits, increased tax credits).<br>Suicide rates increased when governments introduced austerity measures (stricter criteria for state disability benefits).<br>All four studies were high quality. |
| Volkos and Symvoulakis 2021 (17)<br>Systematic review | To identify to what extent the economic crisis of the last decade has influenced mental health issues in daily life, by offering an in-depth analysis of such effect. | General population | n=2 | Retrospective observational analysis | 2013 and 2018 | Greece, 27 European countries, 18 American countries | A worldwide report showed that there was an increase in suicides in men in 27 European and 18 American countries after the 2008 global financial crisis.<br>The overall quality and risk of bias in the studies included in this review were not discussed by the authors. |
| Cairns, 2017 (22)<br>Systematic review | To explore the extent to which area-level socioeconomic disadvantage is associated with inequalities in suicidal behaviour and self-harm in Europe | General population | n=22 | Repeated cross-sectional, prospective and retrospective cohorts, time series, cross-sectional | 2005-2015 | England, Scotland, Northern Ireland, Spain, Republic of Ireland, Finland, Denmark, Sweden, Portugal, Netherlands, Switzerland, and a multi-country study (Slovakia, Italy, Hungary, Sweden, Switzerland and Portugal) | Findings provide strong evidence of increased risk of suicide in areas experiencing high levels of socio-economic disadvantage across Europe.<br>This was consistent across different countries, all age groups and both genders, but was particularly the case for men.<br>All studies included were classified as medium to high quality. |
| Lemmi et al., 2016 (21)<br>Systematic review | To understand the association between suicidal ideations and behaviours and economic poverty in low-income and middle-income countries | General population | n=18 | Ecological, economic modelling, interrupted time series, case-control, cross-sectional | Not reported | WHO regions: Americas, African region, Eastern Mediterranean region, European region, South-East Asia region, and the Western Pacific region | Review investigated the impact of poverty on suicide mortality, drawing from the findings of 18 studies.<br>Across these studies, a total of 31 associations were examined, focusing on various measures of poverty at both the individual and national levels.<br>The majority reported a positive correlation between poverty and suicide mortality.<br>Most studies were of high and acceptable quality (not reported for individual studies). |
| Mucci *et al.*, 2016 (19)<br>Systematic review | To systematically review the impact of the economic crisis on the health of workers | General population | n=4 | Cross-sectional, time series | 2013-2015 | South Korea, Spain, 54 countries (27 in Europe, 18 in America, 8 in Asia, and 1 in Africa), Italy | All four studies included in this review relating to suicide mortality and economic crisis showed that economic crisis and rise in unemployment were associated with increases in suicide.<br>Quality was determined by study type, parameters used and data collection methods.<br>No studies were classified as high quality (not reported for individual studies). |
| Parmar *et al.*, 2016 (18)<br>Systematic review | To systematically identify, critically appraise, and synthesise empirical studies about the impact of the 2008 financial crisis in Europe on health outcomes |  | n=16 | Not reported | 2008-2015 | European wide: Spain, United Kingdom, Greece, 54 countries (27 European), Ireland, United Kingdom (England & Wales), 29 European countries, 8 Western European countries, France, England only, Finland, Iceland | The majority of the 16 studies reported a significant increase in suicides during the 2008 financial crisis.<br>Of the 16 studies included in this review the majority were classified as weak ($n=14$), the remaining two studies were of moderate quality. |
| Oyesanya, Lopez-Morinigo and Dutta, 2015 (20)<br>Systematic review | To provide a systematic update of the evidence concerning the relationship between economic recession and suicide | General population | n=38 | Time series, panel study, cohort | Not reported | United States, Japan, Russia, 22 former Soviet Bloc Countries, South Korea, Sweden, Spain, 27 European Countries, 18 American Countries, 8 Asian Countries One South African Country, Greece, Italy, Europe and North America | 31 studies found a positive association between economic recession and increased suicide rates following the onset of recession.<br>Two studies reported a negative association between economic recession and suicide (i.e., economic recession appearing to be a protective factor for suicide at the ecological level).<br>Two studies did not find an association and three studies were inconclusive in their findings. |
|  |  |  |  |  |  |  | The authors provide a detailed summary of bias/quality assessment but did not use a standardised tool. |
| Rehkopf and Buka, 2006 (23)<br>Systematic review | To synthesise and summarise the current state of evidence regarding variation of suicide rates across local areas with implications for suicide prevention and future research | General population | n=86 | Cross-sectional | 1970-2004 | Europe, Asia, North America and Australia | Among the 221 analyses reported in 86 studies, 45% reported a significant relationship between the socio-economic character of a region and suicide and 55% reported no relation.<br>70% reported an inverse association, such that areas of higher socio-economic position were associated with lower rates of completed suicide.<br>Risk of bias/quality of this study was not reported. |

One review reported a significant association between suicide mortality and debt (15), while a more recent meta-analysis found that the financially stressed were more likely to die by suicide than their non-exposed counterparts (14). All five reviews examining the impact of the 2008 economic recession reported an increase in suicide rates during this period (16–20). One review reported a positive association for both individual and national poverty measures and suicide mortality (21).

Similarly, two reviews reported that areas characterised by high levels of socio-economic disadvantage were at an increased risk of suicide (22,23). A systematic review examining the evidence regarding social security policy reforms in high-income countries found that suicide rates generally declined when measures were introduced to safeguard income (e.g., retirement benefits, increased tax credits) and increased when governments introduced austerity measures (e.g., stricter criteria to qualify for state disability benefits) (24).

### Education (n=3)

Three studies, two random effects meta-analyses (6,13) (see figure 2) and one systematic review (12), found that lower levels of education were associated with suicide in both men and women (6,12,13) (see Table 3). The same studies reported that suicide was associated with a lower degree of educational attainment.

**Table 3:** Characteristics of studies relating to education.

| Review | Aim | Population(s) of interest | No. of studies related to suicide mortality | Types of included studies | Year range of included studies | Countries included | Main Findings |
| --- | --- | --- | --- | --- | --- | --- | --- |
| Qin *et al.*, 2022 (13)<br>Meta-analysis | To provide an overview of research on this topic, to determine the prevalence of socioeconomic, psychiatric, and physical health risk factors in suicide decedents of middle age, and to present the pooled risk of suicide conferred by these factors at midlife | Suicide decedents | n=8 | Cross sectional, cohort | 2003-2017 | New Zealand, Japan, South Korea, Hong Kong, Spain, Sweden, Italy, Norway | Compulsory education only (9 years of education or less) was associated with a 1.24 (95% CI 0.93, 1.66) relative risk of suicide.<br>In a sub-group analysis by gender the relative risk for males was 1.44 (95% CI 1.08, 1.93) and 1.12 (95% CI 0.71, 1.76) for females.<br>All studies relating to education were high quality. |
| Li *et al.*, 2011 (6)<br>Meta-analysis | To systematically review (a) the risk of suicide associated with high prevalence disorders and key SES measures in population-based studies of suicide; and (b) estimates the population attributable risk of suicide associated with psychiatric disorders and SES measures | Not reported | n=4 | Cohort, case-control | 1994-2007 | Denmark, Canada, Sweden, Finland, New Zealand, Lithuania | The pooled relative risk for suicide associated with low (compared to high) educational achievement was 2.18 (95% CI 1.47, 3.22) and 1.46 (95% CI 0.94, 2.34) for males and females, respectively.<br>The pooled risk for suicide associated with level of education (less than secondary schooling) was 2.42 (95% CI 1.03, 5.70) for males and 1.48 (95% CI 0.94, 2.34) for females.<br>The quality of the included studies was not reported. |
| Raschke *et al.*, 2022 (12)<br>Systematic review | To investigate socioeconomic risk factors for suicidal behaviours (suicidal ideation, attempted suicides, and completed suicides) in South Korea | General population | n=3 | Ecological, case-control | 2006-2015 | South Korea | Individuals with lower levels of educational attainment exhibited higher rates of suicide deaths.<br>College education was linked to the least risk of death by suicide in all considered studies. |
|  |  |  |  |  |  |  | • Differences in suicide mortality between the educational groups grew larger over time and were more profound in men than in women.<br>• Risk of bias was not assessed. |

### Unemployment and job insecurity (n=8)

Six meta-analyses (see Figure 2) and two systematic reviews explored the association of unemployment and unemployment benefits on suicide mortality (6,13,14,25–29) (see Table 4). All six studies reported a positive association between risk of suicide and unemployment (6,14,25–27). One review found a negative correlation between unemployment benefits and suicide rates in three out of five primary ecological studies, with more generous unemployment benefits resulting in fewer suicide deaths (28).

**Table 4:** Characteristics of studies relating to unemployment and job insecurity.

| Review | Aim | Population(s) of interest | No. of studies related to suicide mortality | Types of included studies | Year range of included studies | Countries included | Main Findings |
| --- | --- | --- | --- | --- | --- | --- | --- |
| Rolfes and Shor, 2023 (14)<br>Meta-analysis | To determine the suicide risk following unemployment or financial stress | Individuals with mental illness | n=46 | Case-control, cohort | 1990-2019 | Turkey, Denmark, England, New Zealand, US, Sweden, India, Netherlands, Estonia, Germany, Taiwan, Belgium, Finland, Canada, Australia, UK, Iran, Rwanda, Japan | Unemployed persons were 59.0% more likely to die by suicide (95%CI 27.8%, 97.9%) than those in active employment.<br>For studies of the general population, the unemployed were 87.4% more likely to die by suicide (95% CI 50.1%, 134.1%). Subgroup analyses showed no significant differences by sex.<br>The quality of the included studies was not reported. |
| Amiri, 2022 (29)<br>Meta-analysis | To investigate the impact of unemployment on suicidality | General population, elderly, individuals with heroin use disorder, psychiatric patients | n=21 | Cross-sectional, case-control, prospective cohort | 1987-2019 | United States, India, Denmark, New Zealand, Taiwan, Sweden, Italy, France, Bhutan, Netherlands, Canada, Australia, Puerto Rico, South Korea, Germany, United Kingdom, Slovenia, Finland, Israel, South Africa, Iran, Nigeria, Belgium, Japan | Unemployment was associated with an increased odds of suicide mortality (OR 1.87, 95% CI 1.40, 2.50).<br>The authors assessed and discussed the quality of each study under the following domains: selection bias, confounding, data collection and attrition. |
| Qin *et al.*, 2022 (13)<br>Meta-analysis | To provide an overview of research on this topic, to determine the prevalence of socioeconomic, psychiatric, and physical health risk factors in suicide decedents of | Suicide decedents | n=7 | Cross-sectional, cohort | 2003-2018 | New Zealand, Japan, China, South Korea, Denmark, Spain, Ireland, Italy, Norway | Unemployment was associated with a 3.91 (95% CI 2.73, 5.59) relative risk of suicide.<br>In a sub-group analysis by gender the relative risk for males was 4.10 |
|  | middle age, and to present the pooled risk of suicide conferred by these factors at midlife. |  |  |  |  |  | (95% CI 2.20, 8.33) and 3.47 (95% CI 1.95, 6.16) for females.<br>One study was of low quality with the remaining six moderate-high quality. |
| Milner *et al.*, 2014 (26)<br>Meta-analysis | To quantify the effects of adjustment for mental health on the relationship between unemployment and suicide | Not reported | n=5 | Cohort studies | 2000-2012 | Denmark, Sweden | The overall effect of unemployment was associated with a 1.41 relative risk of suicide (95% CI 1.21, 1.60).<br>After adjusting for mental health, the relative risk of suicide following unemployment was reduced by almost 37% (1.15, 95% CI 1.00, 1.30).<br>Mental health was considered a confounder rather than an effect modifier.<br>Studies included in the meta-analysis were deemed to be of high quality. |
| Milner *et al.*, 2013 (25)<br>Meta-analysis | To summarise evidence to date on the effects of duration of unemployment on suicide attempts and death | Unemployed persons | n=6 | Cohort | 2000-2010 | Denmark, Sweden, Finland | The pooled relative risk of suicide associated with longer unemployment (average follow-up time 7.8 years) compared to those currently employed was 1.70 (95% CI 1.22, 2.18).<br>Only cohort studies were included in the meta-analysis as they are of higher quality than other observational study designs. |
| Li *et al.*, 2011 (6)<br>Meta-analysis | To systematically review (a) the risk of suicide associated with high prevalence disorders and key SES measures in population-based studies of suicide; and (b) estimates the population attributable risk of suicide associated with psychiatric disorders and SES measures | Not reported | n=4 | Case-control, cohort | 2000-2003 | Denmark, Canada, Sweden, Finland, New Zealand, Lithuania | The pooled relative risk for suicide associated with unemployment was 1.68 (95% CI 1.11, 2.54) and 1.68 (95% CI 1.09, 2.59) for males and females, respectively.<br>The quality of the included studies was not reported. |
| Virgolino *et al.*, 2022 (27)<br>Systematic review | To review, integrate, and summarise evidence about the association between unemployment and anxiety disorders, mood disorders, and suicidal behaviour, and identify variables affecting this association | General population (aged 15 years and over) | n=93 | Ecological, cross-sectional, case-control, cohort | 1992-2019 | America, Asia, Australia, Europe | • The majority of studies included in the review show a strong positive association between unemployment and suicide mortality.<br>• Evidence provided here demonstrates that men and women respond differently to unemployment.<br>• Risk of suicide was generally higher for unemployed men.<br>• The majority of included studies were medium-high quality (4.8% low). |
| Shand, Duffy and Torok, 2021 (28)<br>Systematic review | To assess whether government responses to unemployment can moderate the impact of unemployment on rates of suicide and self-harm. | General population | n=6 | Ecological, time series | 2009-2017 | Greece, Ireland, Italy, Portugal, Spain, US, EU, North America, Australia, 20 OECD countries | • Based on three of five studies, unemployment benefits were found to be negatively associated with suicide rates.<br>• There is evidence that active unemployment policies and employment protection legislation have beneficial effects.<br>• Policies had a small but meaningful impact on suicide rates in men.<br>• Quality assessment not undertaken. |

### Working life conditions (n=3)

Three random effects meta-analyses (see Figure 2) investigated various components of working life conditions, including job stressors (30), workplace violence (31), workplace bullying (31) and low occupation (6) (see Table 5). One study revealed that exposure to any job stressor was associated with an increased risk of suicide (30). Furthermore, there was a higher risk of suicide associated with lower supervisor and collegial support and low job control. Another study found that the suicide rate was higher in those exposed to workplace violence than among the non-exposed; however, suicide incidence was not significantly higher among individuals exposed to workplace bullying (31). A meta-analysis of the association between occupation and suicide, found that lower skilled occupations (manual/non-skilled/blue collar workers) were at increased risk of suicide than higher skilled occupations. This association was significant among men but not among women (6).

**Table 5:** Characteristics of studies relating to working life conditions.

| Review | Aim | Population(s) of interest | No. of studies related to suicide mortality | Types of included studies | Year range of included studies | Countries included | Main Findings |
| --- | --- | --- | --- | --- | --- | --- | --- |
| Hanson *et al.*, 2023 (31)<br>Meta-analysis | To investigate the association of workplace violence, workplace bullying, and the risk of subsequent suicide attempts or death by suicide | Individuals of working age (15-65 years) | n=4 | Prospective cohort | 1995-2018 | Not reported | • Suicide death was 1.78 (95% CI 1.16, 2.73) times higher in those exposed to workplace violence than in the unexposed.<br>• Similarly, death by suicide was 1.59 times higher in individuals exposed to workplace bullying, though this association was not significant (95% CI 0.97, 2.62).<br>• The studies included in this review were classed as high risk of bias. |
| Milner *et al.*, 2018 (30)<br>Meta-analysis | To investigate whether exposure to job stressors was associated with a greater risk of suicidal ideation and/or behaviours | General population, male sawmill workers, medical school cohort | n=6 | Case-control, cohort | 2007-2017 | Australia, Canada, Germany, Japan | • There was a higher risk of suicide associated with lower supervisor and collegial support and low job control.<br>• There were no significant associations between job demands or job strain and suicide death.<br>• Exposure to any job stressor was associated with a 1.17 (95% CI 1.03 to 1.34) increased risk of suicide.<br>• The quality of the included studies was not reported. |
| Li *et al.*, 2011 (6)<br>Meta-analysis | To systematically review (a) the risk of suicide associated with high prevalence disorders and key SES measures in population-based studies of suicide; and (b) estimates the population attributable risk of suicide associated with psychiatric disorders and SES measures | Not reported | n=3 | Case-control, cohort | 2001-2007 | Denmark, Canada, Sweden, Finland, New Zealand, Lithuania | • The pooled relative risk for suicide associated with low occupation (manual/non-skilled/blue collar workers) was 2.67 (95% CI 1.53, 4.68) and 1.27 (95% CI 0.54, 2.94) for males and females, respectively.<br>• The quality of the included studies was not reported. |

### Housing, basic amenities and the environment (n=21)

Twenty-one reviews (ten meta-analyses (see Figure 3), eleven systematic reviews), covered a broad spectrum of topics, including: rural versus urban living (12,32–35); temperature increase (36–40); air pollution and ozone exposure (39,41–44); natural disasters (45–47); exposure to pesticides (48); and the impact of displacement and housing affordability and foreclosure (49,50) (see Table 6).

**Figure 3:**
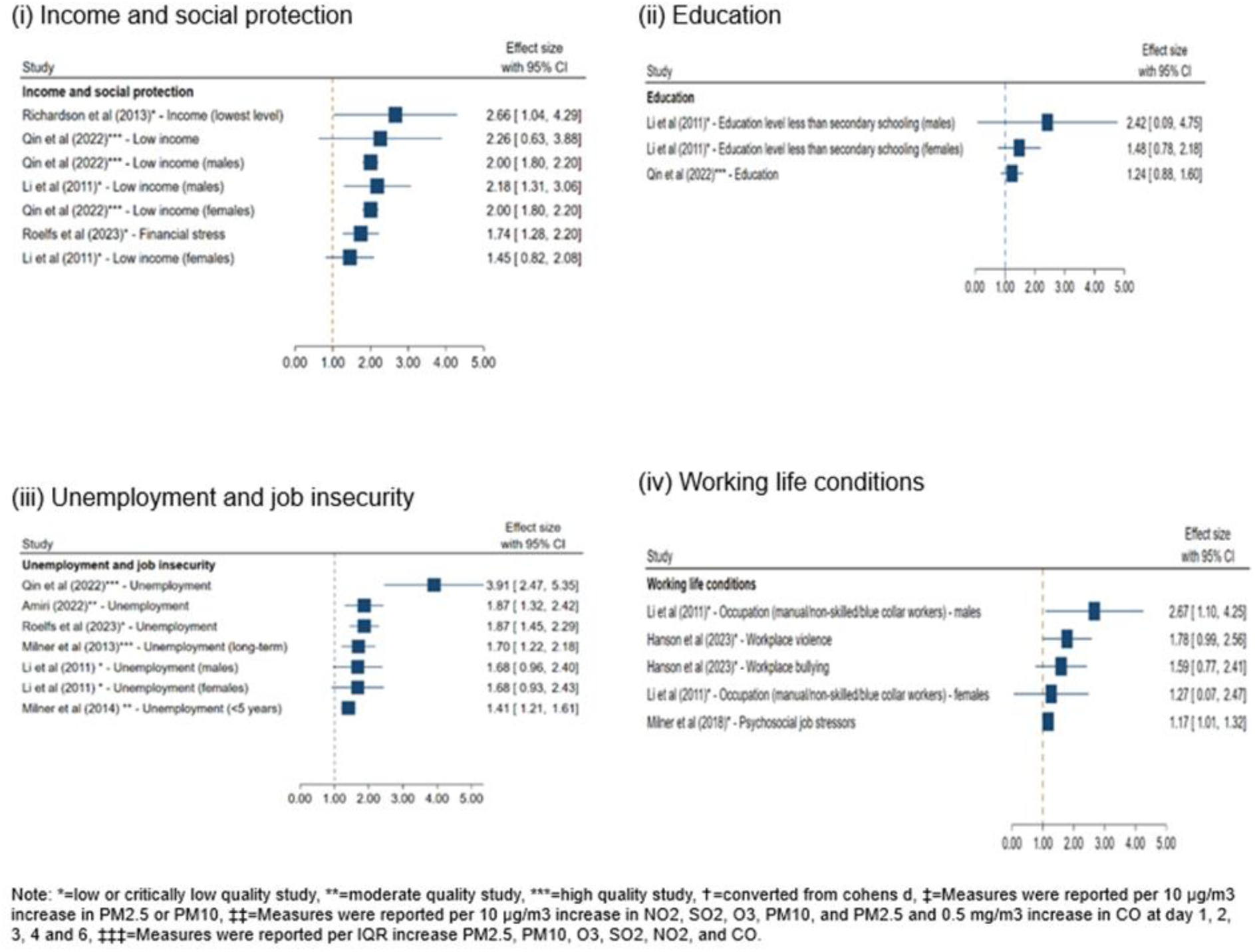
Forest plot visualisation of meta-analyses results for (i) income and social protection; (ii) education; (iii) unemployment and job insecurity and (iv) working life conditions.

**Figure 4:**
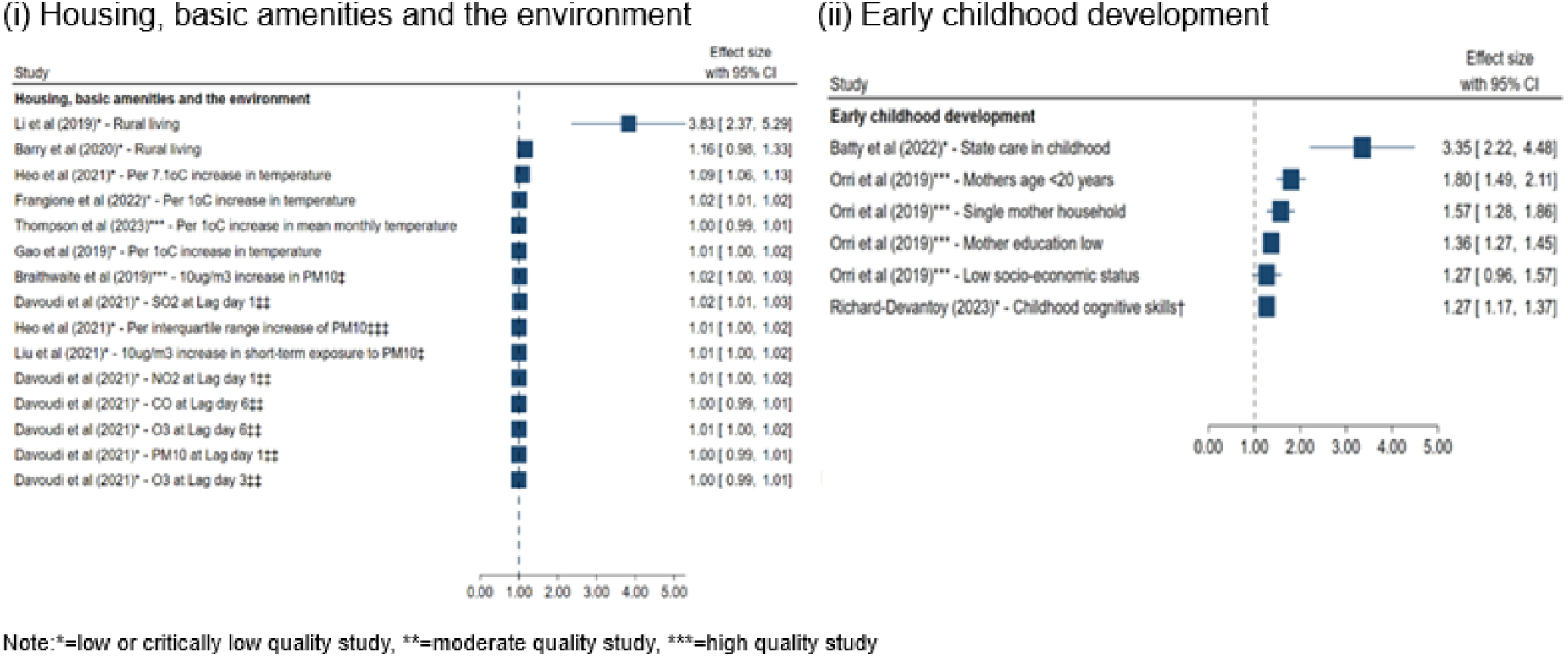
Forest plot visualisation of meta-analyses results for (i) housing, basic amenities and the environment and (ii) early childhood development.

**Table 6:** Characteristics of studies relating to housing basic amenities and the environment.

| Review | Aim | Population(s) of interest | No. of studies related to suicide mortality | Types of included studies | Year range of included studies | Countries included | Main Findings |
| --- | --- | --- | --- | --- | --- | --- | --- |
| Thompson *et al.*, 2023 (36)<br>Meta-analysis | To comprehensively analyse the current evidence regarding the associations between ambient temperature and mental health outcomes | General population | n=9 | Case-crossover | 2011-2021 | United States, Australia, Taiwan, Turkey, Korea, England, Wales | • A 1°C increase in mean monthly temperature was associated with an increase in the incidence of suicide by 1.5% (0.8%, 2.2 %).<br>• A 1°C increase in mean daily temperature was associated with an increase in incidence of 1.7% (0.3%, 3.0%).<br>• A 1°C increase in mean monthly temperature was associated with a risk ratio of 1.01 (95% CI 1.00, 1.01).<br>• Of the studies included, one study was classed as high risk of bias, one moderate risk and the reminding seven as low risk of bias. |
| Frangione *et al.*, 2022 (38)<br>Meta-analysis | To examine the association between either daily, or weekly, variations for eight meteorological variables and suicide outcomes (attempts, or deaths) | General population | n=16 | Time-series, case-crossover, time stratified and case crossover | 1997-2020 | Brazil, United States, Austria, Columbia, Kazakhstan, China, Turkey, 12 countries (not named), Japan, United Kingdom, Spain, Germany, Southern Israel | • Ambient temperature was positively associated with suicide mortality incidence.<br>• In a random effect meta-analysis, the overall relative risk per 1°C increase in temperature was 1.016 (95% CI 1.013, 1.019).<br>• One study included in this meta-analysis was rated “poor”, two “moderate” and the remaining 13 studies were of “high” quality. |
| Davoudi *et al.*, 2021 (41)<br>Meta-analysis | To explore the association between short-term exposure to air pollution and suicide mortality, with an emphasis on different lag times | General population | n=11 | Case-crossover, time series | 2010-2019 | South Korea, United States, China, Japan, Belgium, Colombia, | • Results indicate a positive, though weak, association between air pollution and suicide mortality.<br>• Pollutants such as NO<sub>2</sub>, SO<sub>2</sub>, PM<sub>10</sub>, and PM<sub>2.5</sub>, particularly within the |
|  |  |  |  |  |  | Northeast Asia, Mexico, Taiwan | cumulative lag of 0–1 day, were associated with heightened risks of suicide within the general population.<br>All studies included in the meta-analysis were of moderate to high quality. |
| Heo, Lee and Bell, 2021 (39)<br>Meta-analysis | To investigate suicide risk associated with short-term exposure to **ambient temperature** and air pollution | General population | n=18 | Time series, case crossover | 1994-2019 | Brazil, England, Spain, Austria, USA, Columbia, 29 European countries, Kazakhstan, Finland, South Korea | Relative risk of daily suicide rates per IQR increase in daily temperature was 1.09 (95% CI 1.06, 1.13).<br>14 studies included in the meta-analysis were deemed “probably low risk of bias”, with the remaining four “high risk”. |
| Heo, Lee and Bell, 2021 (39)<br>Meta-analysis | To investigate suicide risk associated with short-term exposure to ambient temperature and **air pollution** | General population | n=13 | Time series, case crossover | 2010-2019 | Mexico, United States, Canada, Colombia, South Korea, Belgium, Asia, China, Japan, Taiwan | Relative risks of suicide per IQR (Interquartile range) increase in PM<sub>2.5</sub>, PM<sub>10</sub>, and NO<sub>2</sub> were 1.02 (95% CI 1.00, 1.05), 1.01 (95% CI 1.00, 1.03), and 1.03 (95% CI 1.00, 1.07).<br>O<sub>3</sub>, SO<sub>2</sub>, and CO were not associated with suicide.<br>Primary studies included in the meta-analysis were classed as “probably low risk” of bias and “definitely low risk” of bias. |
| Liu *et al.*, 2021 (42)<br>Meta-analysis | To determine the overall relationship between PM exposure and depression/suicide | General population | n=10 | Time-series, time stratified case cross-over, cohort | 2010-2019 | Mexico, USA, Belgium, Columbia, Korea, Japan, China, | For a 10 µg/m<sup>3</sup> increase in short-term exposure to PM<sub>2.5</sub>, there was a 2% increased risk of suicide.<br>A 10 µg/m<sup>3</sup> increase in short-term exposure to PM<sub>10</sub> was associated with a 1% increase in suicide risk.<br>Of the studies included in the meta-analysis, three were rated moderate and seven as high quality. |
| Barry *et al.*, 2020 (32) | To determine whether those living in rural areas are more likely to complete or attempt suicide | People living in high-income | n=36 | Prospective cohort, retrospective | 2006-2017 | Canada, United States, United Kingdom, Australia | Living in rural areas was associated with an increased risk of suicide. |
| Meta-analysis |  | English-speaking countries |  | cohort, case control, cross-sectional, ecological |  |  | Gender may moderate this association with rural males being at a greater risk of suicide than urban males.<br>Females may be at a similar risk regardless of residence.<br>12 studies were rated as “high” quality with the remainder moderate-low quality. |
| Safarpour *et al.*, 2020 (45)<br>Meta-analysis | To measure the rate of suicide death after disasters | General population | n=11 | Not reported | 1999-2018 | Japan, USA, Taiwan, China, Sri Lanka | Eleven studies related to suicide death rate before and after disasters estimated suicide death rates of 13.61 per 100,000 people before disasters and 16.68 per 100,000 people after disasters.<br>In a sub-group analysis of five studies according to gender, the suicide death rate before and after disasters was 28.36 and 32.17 among men, and 12.71 and 12.69 among women per 100,000 people, respectively,<br>This indicated an increase of 3.8 suicides among men, and no change for women. |
| Braithwaite *et al.*, 2019 (43)<br>Meta-analysis | To investigate quantitative associations between PM exposure and multiple adverse mental health outcomes | General population | n=4 | Case-crossover | 2010-2017 | United States, Belgium, Republic of Korea, China | The combined effect estimates per 10µg/m<sup>3</sup> PM<sub>10</sub> increment were not statistically significant at lag 0-1 days (RR = 1.01, 95% CI 0.99, 1.03), but they showed significance at lag 0-2 days (RR = 1.02, 95% CI 1.00, 1.03).<br>All four studies included in the meta-analysis were rated as good quality. |
| Gao *et al.*, 2019 (37)<br>Meta-analysis | To investigate if sunlight hours and temperature can affect the risk of suicide | General population | n=11 | Cross-sectional | Not reported | Brazil, Australia, Kazakhstan, Korea, England, Wales, Columbia, Italy, Taiwan, New | A random-effects meta-analysis showed that a 1°C increase in temperature was significantly associated with a 1% increase in the incidence of completed suicide. |
|  |  |  |  |  |  | Zealand, Austria, Greece, Norway | All studies included in the meta-analysis were moderate to high quality. |
| Raschke *et al.*, 2022 (12)<br>Systematic review | To investigate socioeconomic risk factors for suicidal behaviours (suicidal ideation, attempted suicides, and completed suicides) in South Korea | General population | n=3 | Ecological, case control | 2006-2015 | South Korea | Two of the three included studies found that metropolitan residence was associated with lower odds for suicide compared to living in rural or urban areas.<br>Due to the limited number of studies exploring this association, these findings are insufficient to establish definitive conclusions.<br>Risk of bias for the individual studies was not assessed. |
| Satherley *et al.*, 2022 (33)<br>Systematic review | To review the effects of urban living on suicidality and self-harm in the UK and Ireland | Urban residents | n=9 | Ecological | 1960-2019 | United Kingdom, Ireland | Three of nine studies reported a significant association between urban living and suicide (urban areas were more associated with suicide than rural areas).<br>In five of these studies, adjustments were made for area-level socioeconomic deprivation; this socioeconomic deprivation largely accounted for higher rates of suicide in urban areas (3/5 studies).<br>One study reported greater suicide rates in Scottish urban areas with overcrowding and tenement housing.<br>Methodological quality could not be determined for individual studies included in the review. |
| Cogo *et al.*, 2021 (49) | To synthesise what is known about the rates and prevalence of suicide and suicidal behaviour among | Displaced people: refugees granted | n=11 | Monitoring study, cross-sectional, | 2006-2021 | Canada, Denmark, Sweden, Thailand, Netherlands, Sudan | Suicide rates ranged considerably from 4.0 to 290 per 100,000 person-years across the 11 studies. |
| Systematic review | displaced people from published literature. | asylum; refugees in camps; asylum seekers; internally displaced people |  | retrospective cohort |  |  | It was not possible to draw conclusions on the comparative rate of suicide in refugees and host populations.<br>The results from different studies are inconclusive, with several studies suggesting that rates of suicide may be lower in refugees granted asylum than in the host populations.<br>All studies relating to suicide mortality were low quality. |
| Li and Katikireddi, 2019 (35)<br>Systematic Review | To review the incidence of suicide in rural and urban China among the elderly | Elderly (aged 60 or over) living in China | n=7 | Register studies | 1993-2016 | China | Elderly individuals residing in rural regions of China exhibited nearly a fourfold higher likelihood of suicide compared to their urban counterparts.<br>All studies were rated as moderate-low risk of bias. |
| Thompson *et al.*, 2018 (40)<br>Systematic review | To describe the mental health effects of high ambient temperatures and heat waves | General population | n=17 | Time series, case-crossover | 1994-2016 | Switzerland, Austria, USA, Canada, Mexico, Kazakhstan, Finland, Japan, Taiwan, Korea, Belgium, England, Wales, Australia, | 15 of 17 studies found a positive and significant association between increasing temperatures and suicide frequency.<br>Two articles reported but did not quantify a positive relationship.<br>Two articles found no significant relationship between suicide and temperature.<br>Most studies were rated "fair".<br>The quality of many studies was affected by omission and lack of detail on confounding factors. |
| Zhao *et al.*, 2018 (44)<br>Systematic review | To systematically review the epidemiological studies on ambient ozone exposure and mental or behavioural disorders to describe consistent associations as they exist or identify gaps in our current knowledge | General population | n=4 | Case crossover, time series, cross sectional | 2008-2017 | Germany, Taiwan, Korea, Belgium, Canada | Although studies suggest a positive correlation between elevated ozone levels and suicide rates, the low quality of the included studies makes it difficult to establish definitive conclusions. |
| Zhong *et al.*, 2018 (46)<br>Systematic review | To systematically map the long-term health impacts of flooding (including long-term health outcomes, epidemiological trends and impact factors) | General population | n=2 | Mortality case series, prospective cohort | 1996 and 2003 | China, USA | Reported a positive association between flooding and suicide.<br>Methodological quality could not be determined for individual studies included in the review. |
| Downing, 2016 (50)<br>Systematic review | To understand the direct and spillover effect of foreclosures on several health-related outcomes | General population | n=3 | Not reported | 2014-2015 | USA, 20 EU countries | Two studies on suicide at the population-level were included.<br>In a study of 20 European countries, no correlation between unaffordable housing and suicide was found.<br>In a study conducted in the United States, an association between the age-adjusted suicide rate and foreclosures was observed in adults aged 46-64.<br>One study on individual-level foreclosures and suicide reported that the number of foreclosure-related suicides increased during the foreclosure crisis.<br>Risk of bias was not assessed. |
| Fernandez *et al.*, 2015 (47)<br>Systematic review | To systematically map and review available scientific evidence on mental health impacts of floods caused by extended periods of heavy rain in river catchments | General population | n=2 | Longitudinal studies | 1998 and 2013 | USA, Australia | The evidence regarding suicide following a flood event was contradictory.<br>Suicide rates increased following a flood event in the United States, but this finding was not replicated in Australia.<br>No quality assessment tool was used, authors discuss the limitations and biases inherent in longitudinal study design, which may have influenced the findings. |
| Freire and Koifman, 2013 (48)<br>Systematic review | To systematically review epidemiologic literature on the relationship of pesticide exposure with depression and suicide | General population, agricultural workers, rural residents, pesticide applicators and their spouses | n=11 | Ecological, cross-sectional, nested case-control, case-control, prospective cohort | 1996-2011 | Spain, Brazil, USA, New Zealand, Canada, Australia | Four of the eleven studies found that suicide rates were higher in areas with intensive pesticide use compared to areas with lower pesticide use.<br>Risk of bias was not assessed. |
| Clark *et al*, 2007 (34)<br>Systematic review | To assess the strength of the evidence of the impact of the physical environment on mental health and well-being | General population | n=6 | Cross-sectional | 1998-2005 | Australia, USA | Consistent evidence for an association between rural residence in adulthood and suicide rates for males, but not females.<br>The association between rural residence and suicide may relate to a lack of employment opportunities and financial difficulties in rural areas.<br>All studies included in the review were low quality. |

Five reviews found an association between rural residence in adulthood and risk of suicide (12,32–35). Two reviews concluded that there was an association between urban living and suicide (32,33). Raschke *et al*. (2022) reported that residing in metropolitan areas was associated with lower suicide rates and reduced odds of suicide compared to living in rural or urban areas of South Korea (in two of three studies included) (12).

Five reviews, including four meta-analyses, reported positive, albeit weak, associations between temperature increases and suicide mortality (36–40). Four meta-analyses reported a weak association between air pollution and suicide mortality; however, the association varied across different pollutants and lag times (39,41–44). Finally, one review of four observational studies reported a positive association between ozone exposure and suicide(44).

Three reviews, comprising one meta-analysis and two systematic reviews, synthesised findings relating to the association between natural disasters and suicide mortality (45–47). Overall, Safarpour *et al*. (2020) found that suicide deaths increased significantly in the period after disasters; a subgroup analysis of five studies revealed that this impact was attributable to increases among men (45).

Similarly, another review reported a positive association between flooding and suicide (46); however these findings were based on just two primary studies. Fernandez *et al*. (2015) reported conflicting evidence about changes in suicide incidence in the aftermath of a flooding event (47).

One systematic review presented conflicting evidence on the association between pesticide exposure and suicide mortality. Only four out of eleven primary studies reported that suicide rates were higher in areas with intensive pesticide use (48).

In a systematic review investigating suicide rates among displaced populations, several studies suggested that suicide rates were lower in refugees who have been granted asylum compared to the host populations (49).

Finally, a systematic review, including two population-level studies and one individual-level study, synthesised the evidence regarding housing and suicide (50). The first population-level study found no correlation between unaffordable housing and suicide, while the second study reported an association between the age-adjusted suicide rate and foreclosures, particularly among adults aged 46-64 years. The individual-level study reported an increase in foreclosure-related suicides during the foreclosure crisis in 2007.

### Early childhood development (n=6)

Three systematic reviews and three meta-analyses (see Figure 3) investigated various aspects of early childhood development, including: child welfare services (51,52), perinatal exposures (53–55) and cognitive skills (56) (see Table 7). A meta-analysis reported a significant association between adults exposed to any form of state care in childhood and the risk of suicide in adulthood (51). Similarly, another systematic review indicated an increased suicide risk among individuals who have received child welfare services interventions (52).

**Table 7:** Characteristics of studies relating to early childhood development.

| Review | Aim | Population(s) of interest | No. of studies related to suicide mortality | Types of included studies | Year range of included studies | Countries included | Main Findings |
| --- | --- | --- | --- | --- | --- | --- | --- |
| Richard-Devantoy *et al.*, 2023 (56)<br>Meta-analysis | To examine the association between childhood cognitive skills and adult suicidal behaviour, namely attempt and mortality | General population, military conscripts | n=16 (11 in meta-analysis) | Population-based longitudinal studies | 2008-2021 | Sweden, Scotland, Norway, Britain, Israel | Findings indicate that low childhood cognitive skills are associated with increased risk of suicide.<br>Educational attainment was found to be a mediator for this association.<br>All studies were moderate to high quality. |
| Batty *et al.*, 2022 (51)<br>Meta-analysis | To synthesise evidence on the risk of adult mortality in people with a history of state care in early life, and assess the association according to different contexts | Children in state care | n=4 | Prospective cohort | 1995-2018 | Canada, Finland, Sweden | Adults exposed to any form of state care in childhood were found to have more than 3 times the risk of suicide in adult compared to their unexposed counterparts.<br>One study was of high quality while the remaining three were low quality. |
| Orri *et al.*, 2019 (53)<br>Meta-analysis | To investigate in-utero and perinatal exposures associated with suicide, suicide attempt, and suicidal ideation | General population | n=14 | Cohort | 2001-2018 | Europe, North America, Taiwan, New Zealand, Brazil | Family/parental characteristics including having a young mother, single mother household and low maternal and paternal education were associated with higher suicide risk.<br>The authors report that the methodological quality of the included studies was high. |
| Vidal-Ribas *et al.*, 2022 (55)<br>Systematic review | To synthesise evidence on early life vulnerability to suicide beginning in the prenatal period and extending through age 12 | Children aged 12 and under | n=54 | Cohort, nested case-control, case-control | Not reported | Finland, Sweden, Greenland, Norway, Scotland, Taiwan, England, United States, Denmark, United Kingdom, Quebec, | Thirteen studies found that children of mothers less than 20 years of age, had up to a 2.5-fold higher risk of suicide than children of mothers in their late 20s.<br>Four studies did not find an association of younger maternal age with child suicide risk. |
|  |  |  |  |  |  | Copenhagen, Switzerland | • Four studies found children of parents with lower education had 50% higher suicide risk.<br>• Seven studies examining parental occupation and income did not find a correlation with suicide risk, except for one large cohort, in which lower parental occupation was associated with higher suicide risk.<br>• Four studies found that immigrant status was associated with higher suicide risk. Risk of bias in the included studies was low. |
| Milde *et al.*, 2021 (52)<br>Systematic review | To gain updated knowledge on how children and youth who have received or are receiving Child Welfare Service interventions from the Nordic Child Welfare Service fare in relation to suicidality | Children in Nordic Child Welfare Services | n=5 | Cohort | 2003-2018 | Sweden, Finland | • All five of the included articles indicated an increased suicide risk for individuals who have received child welfare services interventions.<br>• The quality of the included papers was not reported. |
| Galobardes, Lynch and Davey Smith, 2004 (54)<br>Systematic review | To examine individual-level studies examining childhood socioeconomic circumstances and adult overall and cause-specific mortality | Adults aged 18 and over | n=2 | Cohort | 1998 and 2003 | United Kingdom, Finland | • Two studies reported separately on associations of childhood socioeconomic position with risk of suicide in adulthood.<br>• Despite only 11 suicides occurring in the 1946 United Kingdom birth cohort, suicide was more likely among those with lower childhood socioeconomic backgrounds.<br>• In the Finnish cohort, men whose fathers worked in manual occupations had a higher risk of suicide death; this was accounted for by adult socioeconomic position.<br>• Parental social class was not associated with suicide in women.<br>• Risk of bias for the included studies was not assessed. |

Two reviews found an association between young parent age at birth (<20 years) and education and the risk of suicide (53,55). In another review, perinatal exposures, such living in single mother household and low maternal SES, were associated with higher suicide risk (53). Another review reported an association between childhood socioeconomic position and risk of suicide in adulthood, with suicide being more likely to occur among those of lower childhood socioeconomic background (54). Similarly, men whose fathers worked in manual occupations were also at an increased risk however, this was accounted for by adult socioeconomic position (54). In contrast, another review failed to find an association between suicide risk and unskilled parental occupation (55).

Finally, a meta-analysis of eleven studies indicated that low childhood cognitive skills, for example low IQ and school performance, were associated with increased risk of suicide, mediated by educational attainment (56).

### Social inclusion and non-discrimination (n=3)

Three systematic reviews explored the relationship between social inclusion and suicide mortality (57–59) (see Table 8). One systematic review presented mixed results on the association between social capital and suicide, with only one of the included studies reporting an association (57). Another systematic review examining the literature on social factors influencing suicidal behaviour in older adults found that limited social connectedness was linked to suicide in later stages of life while living with children was associated with a reduced risk of death by suicide (58).

**Table 8:** Characteristics of studies relating to social inclusion and non-discrimination.

| Review | Aim | Population(s) of interest | No. of studies related to suicide mortality | Types of included studies | Year range of included studies | Countries included | Main Findings |
| --- | --- | --- | --- | --- | --- | --- | --- |
| Blázquez-Fernández *et al.*, 2023 (59)<br>Systematic review | To identify an updated association between isolation and suicides | Mixed (general population, children, adults over 18 years, adolescents) | n=8 | Cohort, case-control, cross-sectional, case series | 2016-2022 | South Korea, Mauritius, UK, USA, China, Swaziland, El Salvador | Five studies reported a positive relationship between social isolation and suicides.<br>Eight studies indicated that loneliness increased the likelihood of suicide.<br>The quality of these studies was not reported. |
| Fässberg *et al.*, 2012 (58)<br>Systematic review | To conduct a systematic analysis of studies with comparison groups that examined the associations between social factors and suicidal behaviour (including ideation, non-fatal suicidal behaviour, or deaths) among individuals aged 65 and older | Adults aged 65 years and over | n=3 (with four publications) | Clinical, psychological autopsy, nested-clinical, population-based | 2001-2005 | United States, Sweden, Hong Kong | Findings limited social connectedness is associated with suicide in later life.<br>Based on one study, living with children was associated with decreased risk for death by suicide.<br>Findings from two studies on residential change over the past two years found no association with suicide.<br>The quality of these studies was not reported. |
| De Silva *et al.*, 2005 (57)<br>Systematic review | To systematically review quantitative studies examining the association between social capital and mental illness | General population | n=3 | Ecological, longitudinal | 2003 | United States | One study reported a higher risk of suicide in areas with higher structural social capital (unpublished data) while the other two reported no association.<br>The authors report that methodological limitations were evaluated, documented and are presented as part of the review. Authors report that all studies had methodological limitations. |

Finally, in their 2023 systematic review, Blázquez-Fernández *et al*. demonstrated a positive and direct relationship between suicide mortality and social isolation and loneliness (59).

## DISCUSSION

This umbrella review of 49 reviews, representing seven (of ten) distinct social determinant domains, examined a wide range of associations with suicide risk.

Overall, there was evidence of a modest effect of social determinants on suicide mortality. In particular, suicide risk was associated with unemployment and job insecurity, income and social protection. In addition, a risk between childhood adversity – particularly experiences of care – and suicide was found. The quality of the included reviews varied considerably, as did the strength of the associations reported in the included meta-analyses. High-quality research which fully explores the relationship between social and environmental factors and suicide risk is needed.

The findings of this review support previous research in identifying a direct association between economic instability and suicide, at both individual and ecological levels. Our review also clearly evidences the effects of economic recessions on suicide incidence, again reflecting previous research on this topic (60–62). The finding that suicide rates are positively influenced by governmental measures to protect income but increase following the imposition of austerity measures (24) clearly evidences the impact of governmental economic policies on suicide prevention (63). This could include initiatives such as increased unemployment benefits, welfare programs or stimulus packages aimed at supporting individuals and families facing financial hardship (64). On the other hand, the imposition of austerity measures, which typically involve reductions in government spending and availability of social services, tends to exacerbate economic distress and may increase suicidal risk (65,66). Strategies which aim to support vulnerable groups (e.g. those at risk of job losses or bankruptcy) during periods of uncertainty and economic and global crises should be implemented (67,68). Further research should consider how impacts differ according to income status of countries (69).

At an individual level, there are well established links between social and health inequalities and suicide risk (70,71), with emerging evidence for the role of several factors, including physical health, sociodemographic factors, early childhood events, and interactions with the criminal justice system (71). Our review found that exposure to childhood adversity, represented by involvement with child welfare services and state care, conferred an increased risk of suicide in adulthood. This was also reflected in studies which examined childhood and parental socioeconomic status. This evidence adds to an increasing body of literature which indicates the need for more upstream approaches to preventing suicide, particularly those which involve primary prevention efforts targeted at young people and families (72).

The majority of included studies (*n*=21) focused on the relationship between environmental factors and suicide, most often, air temperature and pollution, and natural disasters. While the evidence linking these environmental factors to suicide risk was generally weaker and suffered in terms of methodological quality, it underscores an emerging area of concern, namely, the impact of climate change on mental health. Given alarming trends in global temperatures and the increased frequency of natural disasters, understanding the mental health consequences of these environmental changes becomes increasingly important in prevention efforts (73,74).

It has been noted that a challenge with evidencing the role of social determinants on health outcomes is their distal relationship, along with the complex interplay between intermediary factors (75), and may be one reason why the effects observed in this umbrella review are of moderate association. In particular, the association between social determinants and suicide may operate through a range of risk factors, such as substance misuse, relationship difficulties, poor mental and physical health (3,76,77). Findings from this review underscore the importance of longitudinal and high-quality research to disentangle the complexity of interactions between various social determinants and suicide risk. Many of the included studies in each systematic review were cross-sectional in design, limiting the ability to infer causality. Future research should aim to employ longitudinal designs and robust methodologies, utilising data from multiple sources, to improve understanding of the temporal relationships and potential causal pathways between social determinants and suicide (75).

The findings of this review are consistent with other reviews examining social determinants of mental health more broadly. In particular, we note a recent evidence review which hypothesises that inequalities experienced at multiple levels arise via structural processes and policies that further marginalise and place vulnerable groups at a significant disadvantage in society (78). In addition, a recent systematic review of reviews (79) provides evidence to support a range of interventions to reduce the impact of social determinants of mental health, especially interventions targeting intimate partner violence and programmes to address working conditions and unemployment. The authors argue for a move away from traditional ‘Western’ psychosocial approaches towards strategies that reduce the impacts of social determinants, such as climate action and food security in the context of natural disasters (79). We would argue that multilevel approaches to suicide prevention are needed, underpinned by a broader public health, whole-of-government strategy (80).

### Implications for policy, practice and research

The growing recognition of the need to consider the intersectionality between social determinants of health and suicide must be reflected in the future development of suicide prevention strategies. In addition, the impact of governmental policies across several sectors should consider the potential mental health impact of such policies, through the use of mental health impact assessments for example (70,71). It is also important that research considers what policies and policy settings are likely to be the most impactful and cost-effective in terms of reducing suicide at a population level.

The observed association between factors rooted in childhood experiences and environments and suicide risk has significant implications, further emphasising the importance of prioritising strategies to reduce suicidal risk among young people, particularly those focused on clinical, educational and community settings (81,82).

This also has implications for the structure and delivery of healthcare supports and services. The findings of this review underline the need to adequately design and deliver supports and interventions for people at risk of suicide that are culturally sensitive and trauma-informed (83).

Our review did not identify any systematic reviews which examined the impact between access to affordable healthcare and suicide risk. However, it is well established that longstanding challenges exist in ensuring that health services, in particular mental health services, are equally accessible across groups (84). These challenges include addressing barriers such as stigma, acceptability and integration of health services (84).

### Strengths and limitations

This umbrella review used a robust methodology to identify evidence supporting the association between various social determinants and risk of suicide. The classification of social determinants developed by the World Health Organization (1) provides a theoretical underpinning of the search process used in the review. The methodological rigour of the review process enhances the reliability and validity of the findings. There are several factors to consider when interpreting the findings of this review. This review has highlighted the need for more high-quality studies examining the social determinants of suicide mortality. Significant heterogeneity was observed in the exposures examined, even within specific categories of determinants. Furthermore, many social determinants were not represented in this review. Challenges have been highlighted when considering the evidence for social determinants of health more broadly (75), underlying the need for high-quality data. This is particularly true when examining suicide, a relatively rare outcome with well-established challenges around accurate reporting across different countries (85).

Some primary studies are included in multiple systematic reviews and meta-analyses in this umbrella review, particularly those focusing on air pollution and temperature. This may lead to potential biases in the overview if these studies disproportionately influence results. Additionally, we observed substantial heterogeneity in the study populations and range of indicators used, ruling out the feasibility of undertaking a meta-analysis. Instead, we opted for a narrative approach to synthesise and interpret the findings from each review, allowing for a comprehensive exploration of the topic while acknowledging the diversity of evidence within the literature. Moreover, the quality of the included systematic reviews and meta-analyses also varied in the rigor of their methodology, risk of bias assessments, and reporting standards. Therefore, it is imperative to consider the quality and reliability of each included review when interpreting the findings presented here. Most of the reviews we included were rated as being critically low (n=25) or low (n=13), particularly those focusing on the domain of the environment. This raises concerns regarding the robustness and reliability of the findings, necessitating cautious interpretation and consideration of the potential for bias and methodological weaknesses.

While the WHO framework of social determinants provided a useful heuristic for this review, it may not encompass all relevant social determinants that contribute to suicide mortality across different populations and contexts. We were also unable to find operational definitions of the WHO concepts, which had repercussions for the development of the search terms for the review. There is a need for a more expansive and inclusive conceptualisation of social determinants that considers additional dimensions, such as cultural factors, interpersonal relationships, violence and crime. Considering these limitations, it becomes evident that further research is essential to address the gaps and challenges identified in this umbrella review and to further our understanding of the complex interplay between a wide and varying array of social determinants and their influence on suicide mortality.

## CONCLUSION

This review has highlighted that the need for high-quality research which addresses the long-term associations between social determinants of health and suicide, with few studies rigorously addressing determinants such as environmental impacts, food security, structural conflict and access to healthcare. Nevertheless, there was evidence for a range of determinants related to income, social protection, unemployment in addition to early childhood development. These findings emphasise the need to advocate for measures which seek to implement population-based approaches to suicide prevention, and to better understand the impacts of measures which seek to address social determinants in terms of mental health and suicide prevention.

## Data Availability

All data produced in the present work are contained in the manuscript

## ACKNOWLEDGEMENTS SECTION

### 1. Funding statement

This work was funded by the Health Service Executive, National Office for Suicide Prevention, Dublin, Ireland.

### 2. Thank-you’s

The authors thank Ms. Virgina Conrick, Academic Success Librarian, University College Cork, for her valuable time and input into the planning and execution of the search strategy used in this review. We would like to thank Dr Cliodhna O’ Brien, Senior Postdoctoral Researcher at the National Suicide Research

Foundation, for her contribution in the development of the umbrella review protocol.

### 3. Conflict of interest statement

The authors declare that they have no competing interests.

## Notes

### Competing Interest Statement

The authors have declared no competing interest.

### Summary of Updates

Revision to the title of the review.

