## Supplementary File 1 for "The social determinants of suicide: an umbrella review"

**Appendix 1**

**PubMed search strategy and results**

| Search number  (Concept) | Query | Results |
| --- | --- | --- |
| #1  (Suicide) | Suicide [MeSH Terms] OR Suicid* [Title/Abstract] | 117,837 |
| #2  (Access to affordable health services of decent quality) | "Health care facilities, manpower, and services"[MeSH Terms] OR Healthcare* [Title/Abstract] OR "Health Services Accessibility" [MeSH Terms] OR "Health Services Accessibility"[Title/Abstract] OR "Universal Health Care"[MeSH Terms] OR "Universal Health Care"[Title/Abstract] OR "Right to Health"[MeSH Terms] OR "Right to Health"[Title/Abstract] OR "Healthcare Disparities"[MeSH Terms] OR "Healthcare Disparit*"[Title/Abstract] OR "Delivery of Health Care"[MeSH Terms] OR "Delivery of Health Care"[Title/Abstract] OR "Community Health Services"[MeSH Terms] OR "Community Health Services"[Title/Abstract] OR "Health Equit*"[Title/Abstract] OR "Emergency Healthcare"[Title/Abstract] | 4,181,253 |
| #3  (Education) | Education [MeSH Terms] OR Educat*[Title/Abstract] OR "Educational status"[MeSH Terms] OR "Educational status"[Title/Abstract] OR "Literacy"[MeSH Terms] OR "Literacy"[Title/Abstract] OR "Educational level"[Title/Abstract] OR "Educational achievement*"[Title/Abstract] OR "Academic performance"[Title/Abstract] OR "Academic success"[Title/Abstract] | 1,468,653 |
| #4  (Income and social protection) | Income [MeSH Terms] OR Income [Title/Abstract] OR Poverty [MeSH Terms] OR Poverty [Title/Abstract] OR Earning* [Title/Abstract] OR Wage* [Title/Abstract] OR "Income level*"[Title/Abstract] OR "Economic Status"[MeSH Terms] OR "Economic status"[Title/Abstract] OR "Socioeconomic Factors"[MeSH Terms] OR "Socioeconomic Factors"[Title/Abstract] OR "Economic Inequalit*"[Title/Abstract] OR "Economic Security"[Title/Abstract] OR "Social Class"[MeSH Terms] OR "Social Class"[Title/Abstract] | 677,382 |
| #5  (Early childhood development) | "Child Development"[MeSH Terms] OR "Child Development" [Title/Abstract] OR Child [MeSH Terms] OR Child* [Title/Abstract] OR "Language Development"[MeSH Terms] OR "Language Development"[Title/Abstract] OR "Adverse Childhood Experiences" [MeSH Terms] OR "Adverse Childhood Experience*" [Title/Abstract] OR "Adverse Childhood Event*" [Title/Abstract] OR "Early Childhood Development"[Title/Abstract] | 2,781,944 |
| #6  (Food insecurity) | "Food Insecurity"[MeSH Terms] OR "food insecurit*"[Title/Abstract] OR "Food Security"[MeSH Terms] OR "Food Security"[Title/Abstract] OR "healthy food*"[Title/Abstract] OR "Food deprivation"[Title/Abstract] OR "Food poverty"[Title/Abstract] OR "Nutritional deficiency"[Title/Abstract] OR "eating behavi*"[Title/Abstract] OR "Food intake"[Title/Abstract] OR "Hunger"[Title/Abstract] | 106,248 |
| #7  (Structural conflict) | "Social Structure"[MeSH Terms] OR "Structural Conflict"[Title/Abstract] OR "Social Structure"[Title/Abstract] OR "Social protection"[All Fields] OR "Social gradient"[Title/Abstract] OR "Policy"[Title/Abstract] OR "Social Policy"[Title/Abstract] | 284,038 |
| #8  (Social inclusion and non-discrimination) | "Social Inclusion"[MeSH Terms] OR "Social Inclusion"[Title/Abstract] OR "Sociological Factors"[MeSH Terms] OR "Sociological Factors"[Title/Abstract] OR "Social Behavior"[MeSH Terms] OR "social behavi*"[Title/Abstract] OR "Lifestyle"[Title/Abstract] OR "social attitude*"[Title/Abstract] OR "Social Deprivation"[MeSH Terms] OR "Social Deprivation"[Title/Abstract] OR "Social Participation"[MeSH Terms] OR "Social Participation"[Title/Abstract] OR "Social Discrimination"[MeSH Terms] OR "Social Discrimination"[Title/Abstract] OR "Discrimination"[Title/Abstract] OR "Social cohesion"[Title/Abstract] OR "interpersonal relations*"[Title/Abstract] OR "Social Norms"[MeSH Terms] OR "Social Norms"[Title/Abstract] OR "Prejudice"[MeSH Terms] OR "Prejudice"[Title/Abstract] OR "Racism"[MeSH Terms] OR "Racism"[Title/Abstract] OR "Equality"[Title/Abstract] OR "Social status"[Title/Abstract] OR "social environment*"[Title/Abstract] | 1,299,700 |
| #9  (Unemployment and job security) | "Employment"[MeSH Terms] OR "Employment"[Title/Abstract] OR "Unemployment"[MeSH Terms] OR "Unemployment"[Title/Abstract] OR "employment, supported"[MeSH Terms] OR "Employment status"[Title/Abstract] OR "job loss*"[Title/Abstract] OR "Job security"[Title/Abstract] OR "Joblessness"[Title/Abstract] OR "Redundancy"[Title/Abstract] OR "Working status"[Title/Abstract] OR "Employment situation"[Title/Abstract] | 187,810 |
| #10  (Working life conditions) | "Occupational Health"[MeSH Terms] OR "Occupational Health"[Title/Abstract] OR "Occupational Safety"[Title/Abstract] OR "Teleworking"[MeSH Terms] OR "Teleworking"[Title/Abstract] OR "remote work*"[Title/Abstract] OR "Workplace Flexibility"[Title/Abstract] OR "Occupational Exposure"[MeSH Terms] OR "Occupational Exposure"[Title/Abstract] OR "Workplace"[MeSH Terms] OR "Workplace"[Title/Abstract] OR "Occupational stress"[MeSH Terms] OR "Occupational stress"[Title/Abstract] OR "workload"[MeSH Terms] OR "workload*"[Title/Abstract] OR "Work Load"[Title/Abstract] OR "Working Conditions"[MeSH Terms] OR "working condition*"[Title/Abstract] OR "burnout, professional"[MeSH Terms] OR "Burnout"[Title/Abstract] OR "Job Enrichment"[Title/Abstract] OR "employee wel*"[Title/Abstract] OR "Job Demands"[Title/Abstract] OR "Organizational Climate"[Title/Abstract] OR "Quality of Work Life"[Title/Abstract] OR "work related illness*"[Title/Abstract] OR "Work-Life Balance"[Title/Abstract] OR "Work Environment"[Title/Abstract] | 269,529 |
| #11  (Housing, basic amenities and the environment) | "Ill-Housed Persons"[MeSH Terms] OR Homeless* [Title/Abstract] OR Housing [MeSH Terms] OR Housing [Title/Abstract] OR "Medically Underserved Area" [MeSH Terms] OR "Medically Underserved Area" [Title/Abstract] OR "Neighborhood Characteristics" [MeSH Terms] OR "Neighborhood Characteristics" [Title/Abstract] OR "Neighbourhood Characteristics" [Title/Abstract] OR Sanitation [MeSH Terms] OR Sanitation [Title/Abstract] OR Environment [MeSH Terms] OR Environment* [Title/Abstract] | 2,779,620 |
| #12 | #2 OR #3 OR #4 OR #5 OR #6 OR #7 OR #8 OR #9 OR #10 OR #11 | 101,705,49 |
| #13 | "Systematic review*" [Title/Abstract] OR "Meta analy*" [Title/Abstract] | 456,153 |
| #14 | #1 AND #12 AND #13 | 1,744 |

**Appendix 2**
