## Supplementary File 2 for "The social determinants of suicide: an umbrella review"

**Studies in which outcomes for suicide mortality could not be separated for overall results.**

| **Author, Year** | **Title** | **Social Determinant** |
| --- | --- | --- |
| Ati *et* *al*., 2021 | What are the risk factors and protective factors of suicidal behavior in adolescents? A systematic review | Early childhood development |
| Carrasco-Barrios *et* *al*., 2020 | Determinants of suicidality in the European general population: A systematic review and meta-analysis | Early childhood development, social inclusion and non-discrimination, unemployment and job insecurity |
| Gariépy *et* *al*., 2021 | The mental health of young people who are not in education, employment, or training: A systematic review and meta-analysis | Education, unemployment and job insecurity |
| Gelvez-Gafaro *et* *al*., 2022 | Psychosocial risk factors associated with suicide in youth and adolescents: A systematic review | Early childhood development |
| Hu *et* *al*., 2023 | Effects of social support on suicide-related behaviors in patients with severe mental illness: A systematic review and meta-analysis | Social inclusion and non-discrimination |
| Hua *et* *al*., 2019 | A systematic review on the relationship between childhood exposure to external cause parental death, including suicide, on subsequent suicidal behaviour | Early childhood development |
| Kim and Leventhal, 2013 | Bullying and suicide | Early childhood development |
| Kim *et* *al*., 2012 | Differences in incidence of injury between rural and urban children in Canada and the USA: a systematic review. | Housing, basic amenities and the environment, early childhood development |
| Lai *et* *al*., 2017 | Factors Influencing Suicide Behaviours in Immigrant and Ethno-Cultural Minority Groups: A Systematic Review | Social inclusion and non-discrimination |
| Lee *et* *al*., 2023 | Meta-Analysis of Acculturation and Suicide-Related Outcomes: A Test of the Immigrant Paradox | Social inclusion and non-discrimination |
| Liu *et* *al*., 2021 | Is there an association between hot weather and poor mental health outcomes? A systematic review and meta-analysis | Housing basic amenities and the environment |
| Mohatt *et* *al*., 2021 | A Systematic Review of Factors Impacting Suicide Risk Among Rural Adults in the United States | Housing, basic amenities and the environment |
| Moore *et* *al*., 2022 | Investigating the relationship between bullying involvement and self-harmful thoughts and behaviour in young people: A systematic review | Early childhood development |
| Ragguett *et* *al*., 2017 | Air pollution, aeroallergens and suicidality: a review of the effects of air pollution and aeroallergens on suicidal behavior and an exploration of possible mechanisms | Housing, basic amenities and the environment |
| Reed *et* *al*., 2021 | Suicide, Race, and Social Work: A Systematic Review of Protective Factors among African Americans | Social inclusion and non-discrimination |
| Todeshkchuei *et* *al*., 2018 | Psychosocial Factors Associated with Suicidal Behavior among Iranian Women: A Meta-analysis | Education, housing basic amenities and the environment |
| Valente *et* *al*., 2023 | Psychosocial Factors Associated with Suicidal Ideation and Behaviour: A Systematic Review | Income and social protection, early childhood development and social isolation and non-discrimination |
| Vásquez-Vera *et* *al*., 2017 | The threat of home eviction and its effects on health through the equity lens: A systematic review | Housing, basic amenities and the environment, Income and social protection |
| Williamson *et* *al*., 2018 | Occupational moral injury and mental health: Systematic review and meta-analysis | Working life conditions |
| Wu *et* *al*., 2023 | Suicide and suicidality in people exposed to pesticides: A systematic review and meta-analysis | Housing basic amenities and the environment |
| Yıldız, 2018 | Suicide in sexual minority populations: A systematic review of evidence-based studies | Social inclusion and non-discrimination |
| Zanchi *et* *al*., 2023 | Could pesticide exposure be implicated in the high incidence rates of depression, anxiety and suicide in farmers? A systematic review | Housing, basic amenities and the environment |
