## Supplementary File 3 for "The social determinants of suicide: an umbrella review"

**Data Extraction Template**

| **Title of Systematic Review** |
| --- |
| **URL to paper** |
| **Author, year of publication** |
| **Aim of review** |
| **Type of review** |
| **Inclusion/exclusion criteria** |
| **Social determinant(s) examined, and definition(s) used** |
| **Number of included studies** |
| **Types of included studies** |
| **Year range of included studies** |
| **Appraisal tool used / operationalised** |
| **Countries** |
| **Study settings** |
| **Populations of interest** |
| **Age range** |
| **Gender** |
| **Measurement tools used (if applicable)** |
| **Total sample size (or study specific sample sizes for meta analysis)** |
| **Findings (for each SD):** |
| **Total number of events** |
| **Descriptive conclusions** |
| **Effect estimates with 95% CI** |
| **Qualitative findings** |
| **Stratification of evidence** |
| **Risk of bias/quality assessment** |
